# Impact of campaign-style delivery of routine vaccines during Intensified Mission Indradhanush in India: a controlled interrupted time-series analysis

**DOI:** 10.1101/2020.05.01.20087288

**Authors:** Emma Clarke-Deelder, Christian Suharlim, Susmita Chatterjee, Logan Brenzel, Arindam Ray, Jessica Cohen, Margaret McConnell, Stephen C Resch, Nicolas A Menzies

## Abstract

**Introduction:** The world is not on track to achieve the goals for immunization coverage and equity described by the World Health Organization’s Global Vaccine Action Plan. In India, only 62% of children had received a full course of basic vaccines in 2016. We evaluated the Intensified Mission Indradhanush (IMI), a campaign-style intervention to increase routine immunization coverage and equity in India, implemented in 2017-2018.

**Methods:** We conducted a comparative interrupted time-series analysis using monthly district-level data on vaccine doses delivered, comparing districts participating and not participating in IMI. We estimated the impact of IMI on coverage and under-coverage (defined as the proportion of children who were unvaccinated) during the four-month implementation period and in subsequent months.

**Findings:** During implementation, IMI increased delivery of thirteen infant vaccines by between 1.6% (95% CI: −6.4, 10.2%) and 13.8% (3.0%, 25.7%). We did not find evidence of a sustained effect during the 8 months after implementation ended. Over the 12 months from the beginning of implementation, IMI reduced under-coverage of childhood vaccination by between 3.9% (−6.9%, 13.7%) and 35.7% (−7.5%, 77.4%). The largest estimated effects were for the first doses of vaccines against diptheria-tetanus-pertussis and polio.

**Interpretation:** IMI had a substantial impact on infant immunization delivery during implementation, but this effect waned after implementation ended. Our findings suggest that campaign-style interventions can increase routine infant immunization coverage and reach formerly unreached children in the shorter term, but other approaches may be needed for sustained coverage improvements.

**Funding:** Bill & Melinda Gates Foundation.

## Introduction

Despite significant investments in improving immunization coverage, the world is not on track to achieve the goals set by the World Health Organization (WHO)’s Global Vaccine Action Plan for 2011-2020 (1). Global coverage of the third dose of diphtheria-tetanus-pertussis (DTP) vaccine, a measure of routine immunization system performance, stagnated between 2011 and 2018 (2). In India in 2016, only 62% of children received a full course of basic vaccines, and state-level coverage ranged from 35% to 91% (4).

There is little rigorous evidence about how to effectively improve routine immunization coverage (5,6). One widely-applied strategy—“periodic intensification of routine immunization”—adapts techniques from mass immunization campaigns and applies them to the delivery of routine vaccines (7)(8). Mass immunization campaigns have been reported to achieve high vaccine coverage (9) of a single or small number of vaccines, but there is little robust evidence on the effectiveness of campaign-like approaches for delivering the full schedule of routine vaccines. While campaign-like approaches can reach many people, some fraction of these individuals may have been reached by routine services anyway. Delivery volumes reported by campaigns may therefore overestimate the net change in coverage.

In 2017-2018, the Government of India implemented a nationwide effort to improve routine immunization coverage and equity, via a campaign-like initiative called Intensified Mission Indradhanush (IMI). IMI was designed to (i) increase coverage of routine vaccines for infants under two and pregnant women in selected low-performing districts, and (ii) sustain these gains by raising public awareness of routine immunization and strengthening routine planning. IMI was one of the largest ever applications of the “periodic intensification of routine immunization” strategy.

In this study, we conducted a quasi-experimental evaluation of IMI. Using a controlled interrupted time-series approach, we estimated the impact of IMI on vaccine delivery, coverage, and under-coverage for 15 vaccines in the routine immunization schedule.

## Methods

### Study setting

India’s immunization program serves a target population of approximately 27 million newborns and 30 million pregnant women annually (10). Vaccinations are provided through public health facilities and community-based outreach sessions, with an estimated nine million immunization sessions held annually (11). The routine schedule includes 25 childhood vaccines and two vaccines for pregnant women (Table 1). Vaccines are administered according to a schedule by age, but some vaccines can be administered late (12).

**Table 1:**
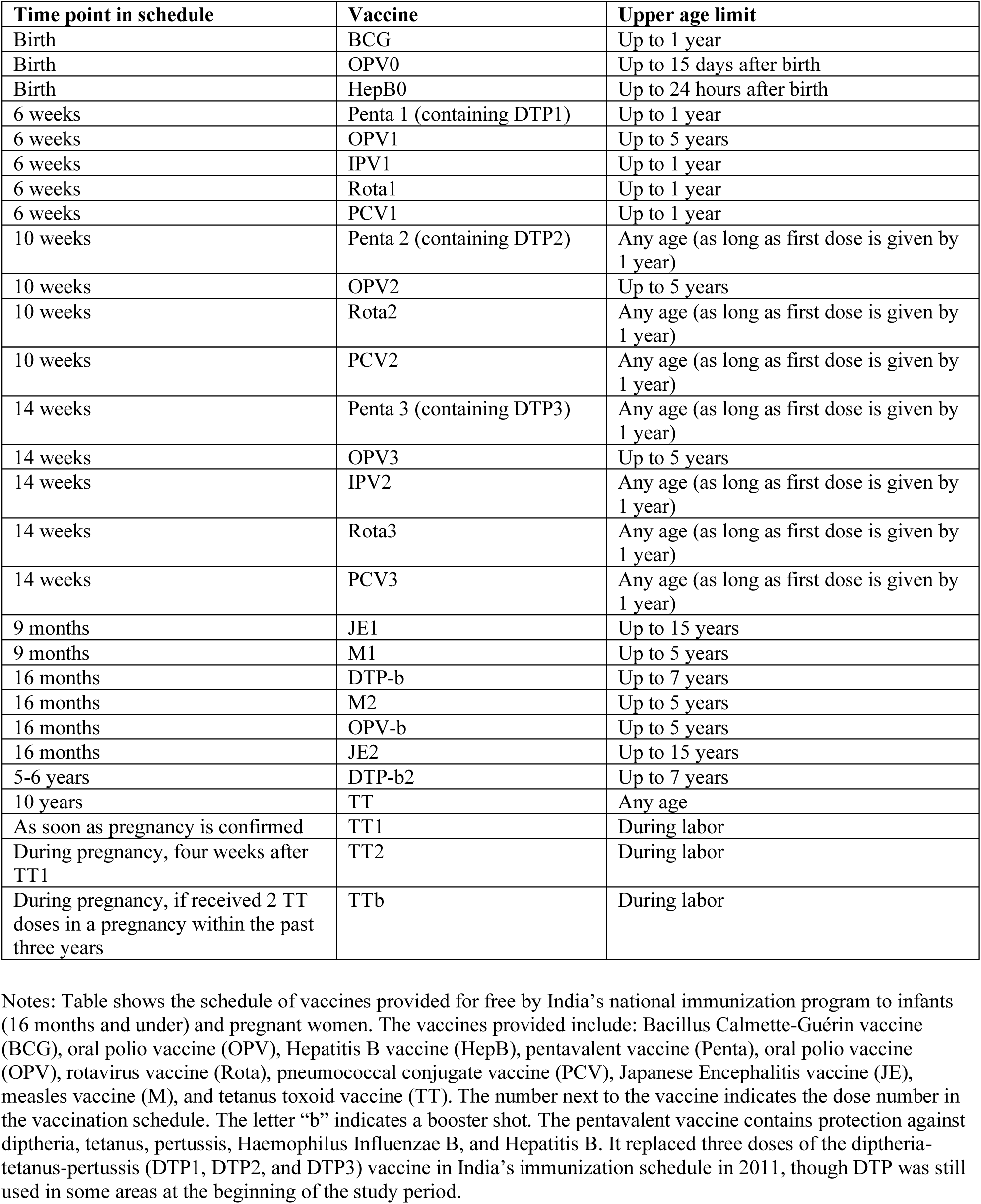
National Immunization Schedule for Children and Pregnant Women in India

Many children and pregnant women do not receive the full schedule of vaccines. Reasons for under-immunization include low awareness (13), inaccessibility of vaccination services (13), anti-vaccine sentiment (14), and under-staffing of health facilities (15). In recent years, in an effort to increase coverage, the Government of India implemented a series of campaign-like interventions called Mission Indradhanush (MI). These interventions fell short of their objectives, leading to the design and implementation of IMI (10).

### Intervention

The Government of India implemented IMI from October 2017 through January 2018 as part of the Pro-Active Governance and Timely Implementation (PRAGATI) initiative, a set of programs prioritized by the office of the Prime Minister. Districts with weak immunization performance (<70% estimated DTP3 coverage, or >13,000 children missing DTP3 in the previous year) were included, and additional districts added based on requests from states (10). In total 187 districts and urban areas were included.

IMI implementation began with door-to-door surveys to identify under-immunized children. District-level micro-plans were then developed to determine the location of IMI vaccination sites and ensure supply availability. Site selection focused on areas with low coverage, with particular emphasis on urban slums and nomadic populations. Social mobilization campaigns were conducted to raise awareness. Finally, immunization sessions were conducted for seven consecutive days per month during implementation. Sessions were conducted by auxiliary nurse-midwives (ANMs), who left their postings in periphery health facilities to deliver vaccines and other health services at the selected locations.

### Data sources

Outcome data at the district-month level were extracted from India’s Health Management Information System (HMIS), which compiles service delivery data reported by health facilities. We used data on vaccine doses delivered from October 2015 through September 2018, encompassing two years before the start of IMI and one year after. We used data on thirteen vaccines for children and two for pregnant women: Hepatitis B birth dose (HepB0), Bacillus-Calmette-Guérin (BCG), four doses of diptheria, tetanus, and pertussis-containing vaccines (DTP1, DTP2, DTP3, and DTP booster (DTPb)), five doses of oral polio vaccine (0PV0, OPV1, OPV2, OPV3, and OPV booster (OPVb)), two doses of measles-containing vaccines (M1 and M2); and the first and second dose (or booster) of tetanus toxoid vaccine (TT1 and TT2) for pregnant women. We also used HMIS data on the number of immunization sessions held per district-month. We excluded the Japanese encephalitis vaccine because it is only delivered in endemic areas. We excluded rotavirus, pneumococcal, and inactivated polio vaccines because they were recently introduced and not available for the full study period. We excluded vaccines for children over age two (a second DTP booster and tetanus toxoid), because IMI primarily targeted children under two and pregnant women. While we included HepB0 and OPV0, we did not expect large effects for these vaccines because the catch-up period is limited to 24 hours and 15 days post-birth, respectively.

To identify districts included in IMI we used publicly available documents from the Indian Universal Immunization Program (16). We also extracted covariate data from India’s 2015-2016 Demographic and Health Surveys (DHS), summarized at the district level (4). This included vaccine coverage and urbanization (percent of children under five living in an urban area). Finally, we used World Bank estimates of India’s 2017 population size, fertility rate, and neonatal mortality rate to generate estimates of the target population size for different vaccines (17).

### Sample

The study sample included all districts in India meeting two criteria: (1) they had available HMIS data for the full study period, and (2) these data could be merged with DHS data. We omitted 8 of 187 IMI districts and 110 of 549 control districts that did not meet these criteria(Table S1).

### Statistical analysis

We conducted a comparative interrupted time-series (CITS) analysis. This quasi-experimental method accounts for time trends that would have occurred in the absence of the intervention, and for exogenous shocks that could have affected the outcome trend in both the treatment and control group (18,19). Our analysis assumed that, in the absence of IMI, deviations from past trends in vaccine delivery would have been the same for treated and control districts.

We modelled time trends in district-level vaccination volume (doses delivered) using generalized linear models with quasi-Poisson distributed outcomes and a log link, in order to appropriately model count data with over-dispersion (20,21). We included dummy variables to model intercept changes on 1 October 2017 (the start of IMI), 1 February 2018, and 1 June 2018. We therefore measured the impact of IMI on vaccine delivery during the four-month implementation period and two four-month post-implementation periods. By including two post-implementation periods, we assessed whether immunization delivery volumes returned to or dipped below their pre-intervention levels after IMI implementation ended. From this analysis we estimated the impact of IMI during the 4-month implementation period, as well as the net impact over the full year following the beginning of implementation.

We assessed whether IMI was more effective in districts with lower coverage or higher levels of urbanization (because these were focus areas of the program) by including interaction terms in our regression models. We also included calendar month fixed effects to adjust for seasonality, and district fixed effects to absorb variation in the initial level of vaccination volume. We used Newey-West standard errors to adjust for potential serial autocorrelation (22).

Because the treatment effect in CITS is captured by multiple coefficients, it is common practice to generate interpretable results by making predictions from fitted models (23). Following this practice, we estimated vaccination volume in the treated districts if IMI had not occurred and compared this to the observed vaccination volume under IMI. We estimated the incremental vaccination volume attributable to IMI by projecting model results to the full set of treated districts with covariates fixed to their true values. To describe how the estimated treatment effect varied by district characteristics, we generated treated effect estimates from models fixing the values of covariates to their 25_th_ and 75_th_ percentiles in the treated districts. We estimated equal-tailed 95% confidence intervals (95% CI) for all effects (24).

To estimate the impact of IMI on coverage in the treated districts, we divided our estimates of incremental doses delivered by estimates of the target population size. We estimated the target population size for HepB, OPV0, BCG, TT1, and TT2 as the number of live births in 2017, calculated as fertility rate multiplied by population size. We estimated the target population size for DTP1-3, OPV1-3, and M1-2 as the number of surviving infants in 2017, calculated by multiplying live births by one minus the neonatal mortality rate. To estimate the impact of IMI on under-coverage (percent reduction in unvaccinated children) we divided our estimates of incremental doses delivered by estimates of the number of children who would not have been reached if IMI had not occurred. We estimated the number of unreached children by multiplying the target population size by one minus coverage in 2016.

To examine mechanisms for the treatment effect, we measured the impact of IMI on the number of immunization sessions held (including both routine and IMI sessions), and the ratio of incremental doses delivered to incremental immunization sessions held.

### Sensitivity analysis

We examined the robustness of our results to different model specifications including single interrupted time-series analysis, a matched analysis using a subset of the study sample (coarsened exact matching on geographic location, baseline coverage, urbanization level, and participation in the earlier MI program), and restriction of the comparison group to untreated districts that do not share borders with treated districts.

Supplementary Appendix A provides detailed analytic methods.

## Results

### Impact of IMI on vaccination volume

During the four-month implementation period, IMI had a positive estimated impact on delivery volume for all infant vaccines in the study (Figures 1 and 2). During implementation, IMI increased infant vaccination volume by between 1.6% (HepB0; 95% CI: −6.4, 10.2) and 13.8 (DTPb; 3.0, 25.7), with a median of 10.6%. There was no statistically discernable impact of IMI on the delivery of vaccines for pregnant women (median −0.9%).

**Figure 1:**
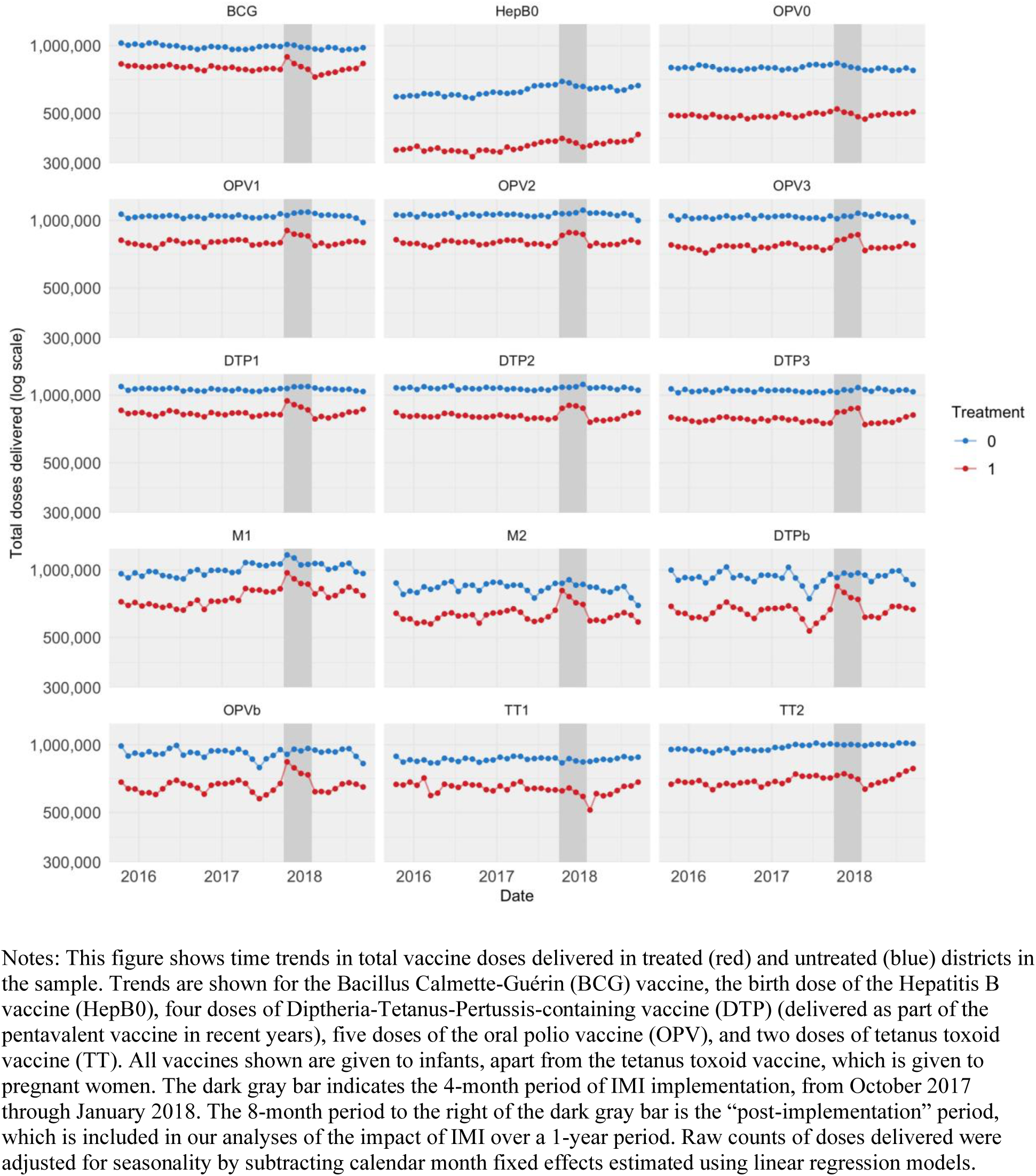
Time trends in vaccination volume in treated and untreated districts, 2 years before and 1 year after the start of IMI implementation

**Figure 2:**
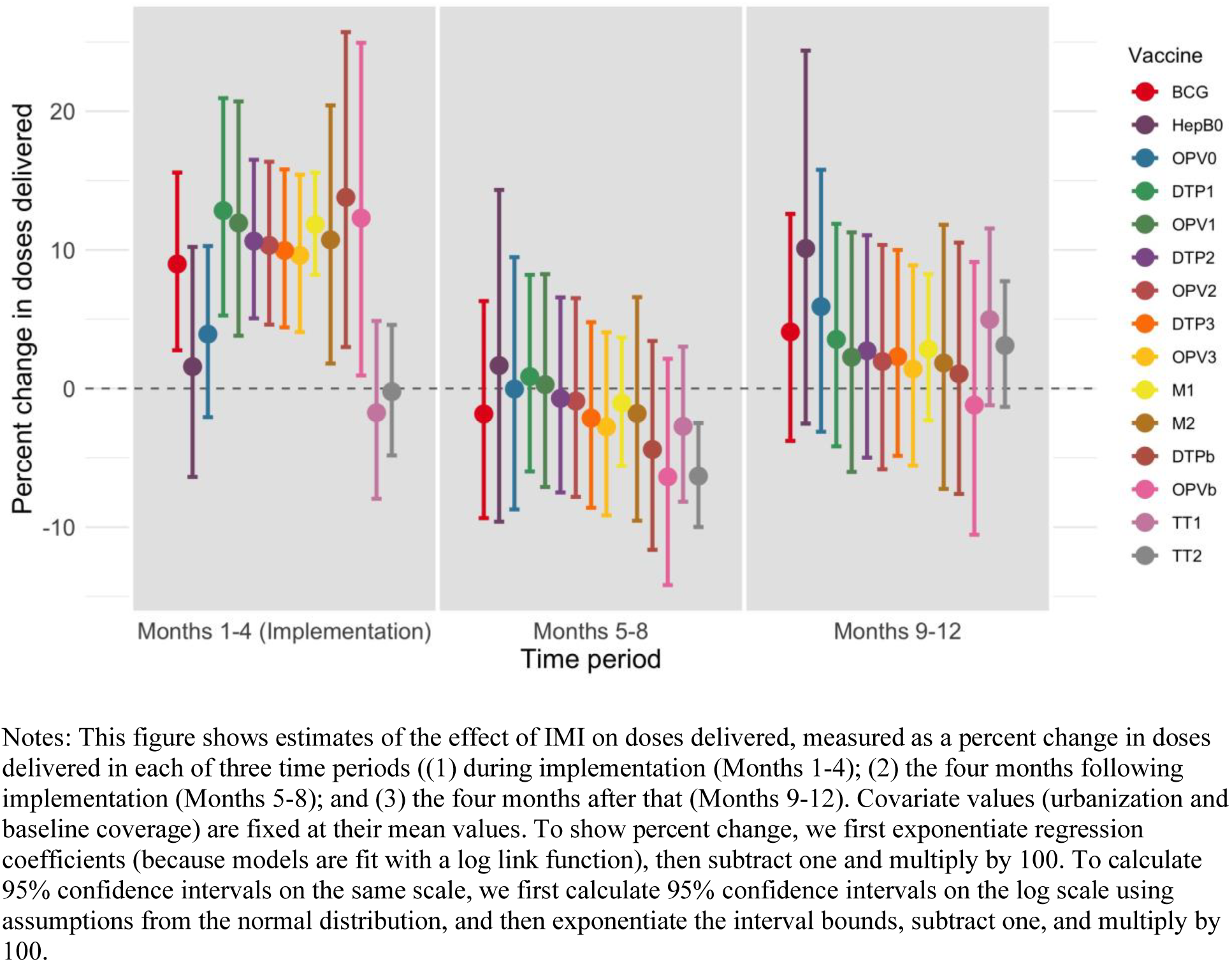
Regression results: percent change in doses delivered in treated districts during three four-month periods

**Figure 3:**
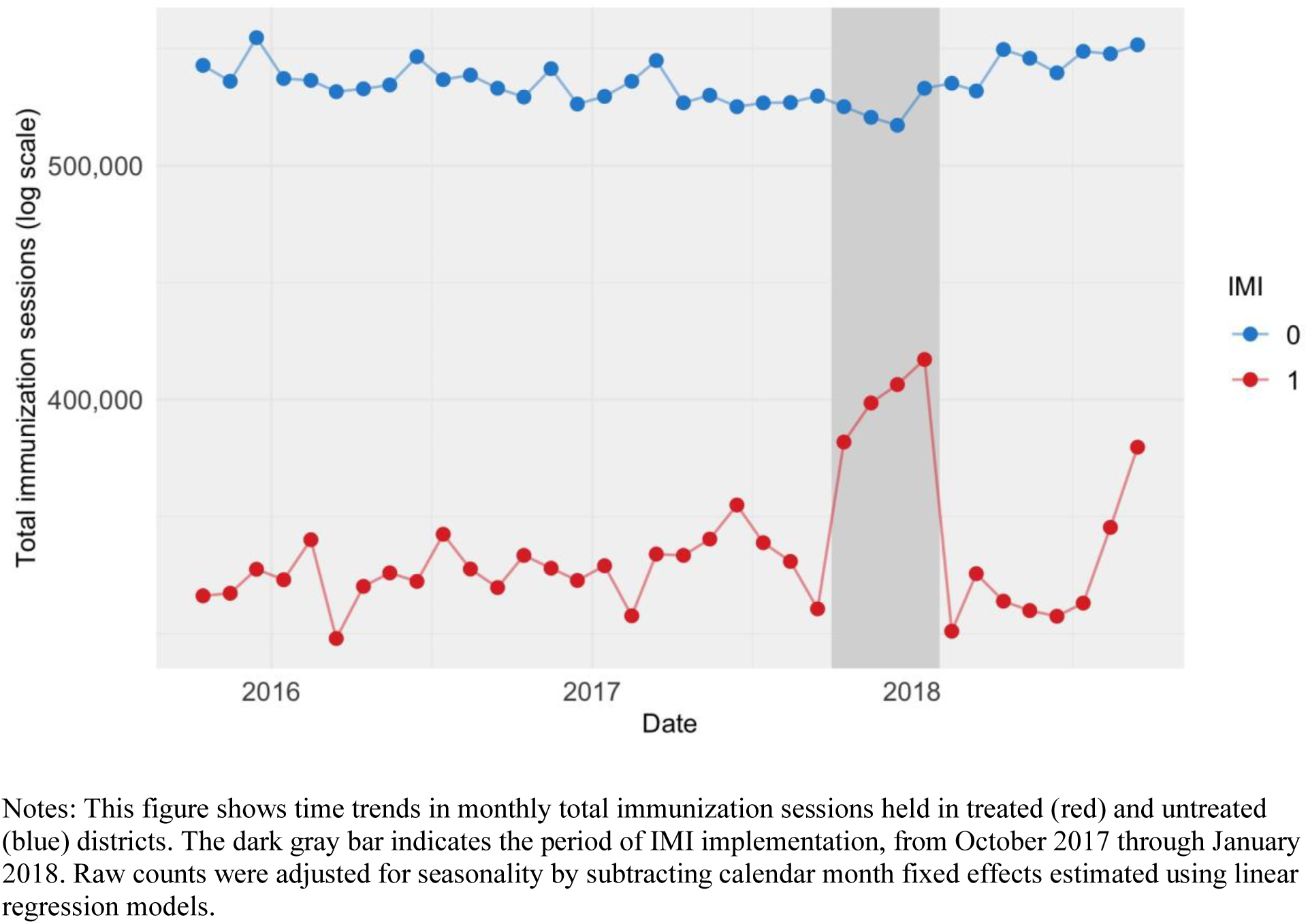
Increase in immunization sessions held during IMI implementation

The estimated effect of IMI waned after implementation ended. With the exception of TT2, there was no statistically discernable impact of IMI on delivery of any antigen during the period from February to May 2018 or the period from June to September 2018. Point estimates for all estimated effects were modestly negative from February to May 2018, with the exception of OPV0, DTP1, and OPV1 (median value −1.8%). Point estimates for all estimated effects were positive from June to September 2018, with the exception of OPVb, but none were statistically significant (median value 2.7%).

Over the full year from the beginning of implementation, we estimated positive impacts of IMI on the number of doses delivered of all infant vaccines and TT1, and a negative impact on doses delivered of TT2 (Table 3). None of these estimates were statistically significant. We estimated that IMI reached between 148,000 (95% CI: −263,000; 521,000) and 491,000 (−100,000; 1,033,000) additional infants with each infant vaccine in the study. We estimated that IMI reached 6,000 (−95% CI: 393,000; 398,000) additional pregnant women with TT1 and −102,000 (−441,000; 215,000) additional pregnant women with TT2.

**Table 3:** Estimated impact of IMI on number of children reached, coverage in the treated districts, and under-coverage in India over one year (October 2017 through September 2018)

|  | (1) Number of additional children reached (thousands) (95% CI) | (2) Percentage point increase in coverage in treated districts (95% CI) | (3) Percentage of unreached children in the treated districts reached through IMI (95% CI) |
| --- | --- | --- | --- |
| BCG | 331<br>(-282; 883) | 3.9<br>(-3.3, 10.4) | 31.5<br>(-26.8, 83.9) |
| HepB0 | 148<br>(-263, 521) | 1.7<br>(-3.1, 6.1) | 3.9<br>(-6.9, 13.7) |
| OPV0 | 136<br>(-299, 500) | 1.6<br>(-3.5, 5.9) | 5.2<br>(-11.5, 19.2) |
| DTP1 | 491<br>(-100; 1,033) | 5.9<br>(-1.2, 12.5) | 35.7<br>(-7.5, 77.4) |
| OPV1 | 396<br>(-221, 969) | 4.8<br>(-2.7, 11.7) | 33.1<br>(-18.5, 81.2) |
| DTP2 | 342<br>(-187; 832) | 4.1<br>(-2.3, 10.1) | 19.7<br>(-10.7, 47.8) |
| OPV2 | 287<br>(-252, 823) | 3.5<br>(-3.0, 9.9) | 17.2<br>(-15.1, 49.4) |
| DTP3 | 253<br>(-274; 733) | 3.1<br>(-3.3, 8.8) | 10.0<br>(-10.9, 29.0) |
| OPV3 | 192<br>(-318, 670) | 2.3<br>(-3.8, 8.1) | 6.9<br>(-11.5, 24.3) |
| M1 | 377<br>(-23; 771) | 4.5<br>(-0.3, 9.3) | 16.9<br>(-1.0, 34.7) |
| DTPb* | 233<br>(-474, 857) | 2.8<br>(-5.7, 10.3) | NA |
| M2* | 247<br>(-372; 793) | 3.0<br>(-4.5, 9.6) | NA |
| OPVb* | 79<br>(-662, 773) | 1.0<br>(-8.0, 9.3) | NA |
| TT1 | 6<br>(-393; 398) | 0.1<br>(-4.6, 4.7) | 0.7<br>(-47.2, 49.9) |
| TT2 | -102<br>(-441; 215) | -1.2<br>(-5.2, 2.5) | -6.5<br>(-27.8, 13.5) |

Notes: Table shows point estimates and 95% confidence intervals reflecting estimation uncertainty. Column 1 shows the estimated number of incremental doses delivered due to IMI. This result is calculated by generating predictions from regression models, with covariates set to their true values in the full set of IMI districts. Column 2 shows the estimated percentage point change in coverage in IMI districts. This result is calculated dividing the values in column 1 by an estimate of the target population. The target population is estimated using World Bank data on the population size, birth rate, and neonatal mortality rate in 2017, and HMIS data on the percentage of live births in India that occurred in IMI treatment districts. The full birth cohort is used as the target population for BCG, TT1, and TT2; the birth cohort size less the infant mortality rate is used as the target population for other infant vaccines. Column 3 shows the estimated percentage of unreached children in the treated districts who were reached through IMI. This result is generated by dividing the values in column 1 by an estimate of the number of children in the target districts who would have been unvaccinated if IMI had not occurred. The denominator is calculated by multiplying the rate of under-coverage in 2016 (from DHS estimates) by the size of the target population. NA indicates “Not Applicable.” \*For these vaccines, there are no estimates of coverage in the 2016 DHS survey.

### Impact of IMI on coverage and under-coverage

Based on these changes in vaccination volume, we estimated percentage point improvements in coverage ranging from −1.2 (−5.2; 2.5) for TT2 to 5.9 (−1.2; 12.5) for DTP1, assessed over the full year following the beginning of implementation.

We estimated that IMI reduced the number of unvaccinated children by between 3.9% (95% CI: −6.9, 13.7) for HepB0 and 35.7% (−7.4; 77.4) for DTP1, and reduced the number of unvaccinated pregnant women by 0.7% for TT1 (−47.2; 49.9) and −6.5% for TT2 (−27.8; 13.5).

### Effects of urbanization and baseline coverage

We found small and inconsistent results for the impact of urbanicity and baseline DTP3 coverage on IMI impact estimates. The estimated impact of IMI on coverage was higher in more urbanized areas for four of the vaccines evaluated, and lower for eleven vaccines. For baseline DTP3 coverage, the estimated impact of IMI was higher in districts with higher baseline coverage for nine of the vaccines evaluated, and lower for five vaccines. Detailed results are provided in Table S2.

### Immunization sessions held, and doses per session

During implementation, IMI was estimated to have increased the number of immunization sessions held per treated district by 11.2% (95% CI: 3.7; 19.4). For the two subsequent four-month periods the number of immunization sessions was estimated to be lower than in the absence of IMI, 7.7% (95% CI: −1.4; 15.9) and 6.1% (95% CI: −4.4; 15.5) lower, respectively. Assessed over the full year period, IMI was estimated to have produced minimal changes in the number of immunization sessions (decrease of 61,000 (95% CI: −325,000; 475,000).

During IMI implementation, the ratio of incremental infant vaccine doses to incremental immunization sessions held ranged from 0.1 (95% CI: −0.9, 0.9) doses of HepB0 per session to 2.9 (1.0; 6.2) doses of DTP per session, and for maternal vaccines the ratio was −0.6 (−8.7, 6.2) doses per session of TT1 and −0.2 (−1.3, 0.8) doses of TT2.

### Sensitivity analyses

We found qualitatively similar results across the eight model specifications we tested, with some variation in point estimates (Figure S6). Detailed results from the sensitivity analyses are included in the Supplementary Appendix (Tables S6-S12).

## Discussion

IMI represents one of the largest efforts to improve routine immunization coverage using campaign methods that has ever been attempted. The focus of IMI was to reach children and pregnant women previously unreached by the vaccination program, to increase coverage and improve equity. Evidence on the effectiveness of this approach can be used to guide future investments in similar types of approaches in India and elsewhere.

We found that IMI substantially increased delivery for thirteen infant vaccines, but not for two vaccines for pregnant women. Assessed over a full year, IMI increased infant vaccine coverage by between 1.6 (−3.5, 5.9) and 4.8 (−2.7, 11.7) percentage points across different vaccines, reaching between 3.9% (−6.9, 13.7) and 35.7% (−7.5, 77.4) of children who otherwise would not have been reached. The largest estimated effects were for DTP1 and OPV1, which suggests that IMI was able to reach previously unvaccinated (“zero dose”) children. The smallest effects were for HepB0 and OPV0; this was expected because these vaccines are only administered up to 24 hours and 15 days after birth, respectively.

Previous RCTs that evaluated interventions similar to IMI, involving immunization outreach at sites closer to communities and social mobilization to increase awareness of vaccination services, found larger effects on coverage (5,25). However, interventions that are successful in the context of an RCT do not always have the same effect when implemented at scale. Outside of a trial setting, the intervention may not be as tailored to the population’s needs, and adherence to program design may not be as consistent. The smaller effect of IMI could also be due in part to low efficiency: while the number of immunization sessions significantly increased during IMI implementation, the efficiency of these sessions (measured as doses delivered per session) was low. We estimated that for infant vaccines, between 0.9 and 2.2 additional doses were delivered per additional session held during the implementation period. This could be because demand was low or because the sessions were not well planned.

An earlier evaluation of IMI also estimated larger (though directionally similar) effects (10). The earlier study used data from household surveys, which have several advantages over the HMIS data used in our study, and can provide direct estimates of coverage. However, this prior study relied on baseline data from two years before the start of IMI and was not able to control for time trends in vaccine delivery that would have occurred in the absence of IMI. This approach would lead to overestimates of the impact of IMI if there were secular improvements in coverage over the two-year period not attributable to IMI. In addition, the prior study did not assess the potential for a rebound effect, whereby vaccination delivery volume decreased in the months following IMI implementation. Our results suggest that evaluations of campaign-style interventions should include a sufficiently long post-period to account for the possibility of rebound after implementation.

While one of the objectives of the IMI program was to have a sustained effect (by raising awareness of the routine immunization program and improving routine planning), we did not find evidence for a sustained effect during the 8-months after IMI implementation ended. During the first four months after implementation, we found a small rebound effect: vaccination volume decreased, though not by a statistically significant amount. During the subsequent four months, vaccination volume increased again, though again not by a statistically significant amount. The presence of a rebound directly after implementation suggests that, for some children, IMI may have improved vaccination timeliness. Reducing delays in immunization can improve health outcomes by to reducing exposed time, though this was not IMI’s objective (26). The lack of evidence for a sustained effect on coverage may be because IMI did not address long-term health system gaps such as human resource constraints and last-mile supply chain challenges. The reasons for lack of sustained impact need further study to inform the design of future interventions.

Although IMI had a special focus on reaching children in urban slum areas and in the poorest performing districts, we did not find substantial variation in treatment effect size by urbanization or baseline coverage (16). Differences in treatment effect size by urbanization and baseline coverage were mostly small and statistically insignificant. This could be driven by our use of district-level data, as it is possible that there was more meaningful variation by urbanization and coverage within districts. It is important to note that all districts included in IMI were selected for weak immunization performance, so, by increasing coverage in the treated districts, IMI had a meaningful impact on geographic equity in India even if the treatment effect size was similar across treated districts.

Our study has several limitations. First, because HMIS data are noisy, our estimates have low precision. We used a quasi-Poisson distribution to accurately estimate uncertainty in the presence of over-dispersion in HMIS data. Second, routinely collected data, such as HMIS data, can be subject to over- or under-reporting. Misreporting that was random, consistent over time, or followed similar trends in treated and untreated districts would not bias our study findings. However, our findings could be biased if IMI influenced reporting practices. While we cannot know for sure, we do not believe this happened: IMI doses were reported into the HMIS system the same way as other doses, and reward payments for health workers did not change during IMI. Third, our main analysis used data on vaccination volume rather than direct measures of coverage. We therefore used auxiliary data sources to calculate target population size for estimating impacts on coverage and under-coverage. The accuracy of our impact estimates for coverage and under-coverage relies on the accuracy of these auxiliary data. Fourth, our causal inference approach relied on the untestable assumption that vaccine delivery trends would have changed the same way in treated and untreated districts if IMI had not occurred. We tested the robustness of our results to a wide range of model specifications and comparison groups, and found qualitatively similar results. Fifth, there were changes in the delivery of the measles vaccine in India that coincided with the study period. As a result, our estimates of the impact of IMI on delivery of M1 and M2 may be less reliable than our estimates for other vaccines. Sixth, our analysis could be strengthened through the use of data on vaccine supply stock and flow, as collected in the electronic vaccine intelligence network (eVIN), but we did not have access to these data. Finally, we did not evaluate the impact of IMI on childhood vaccines delivered after two years, because the target population was under two years, but it is possible that IMI also had an impact on vaccine delivery to the older age group.

### Conclusions

IMI was a major effort to improve immunization coverage in India. We estimate that it had a considerable impact on infant vaccine coverage during implementation, and minimal impacts after this period. “Periodic intensification of routine immunization” may be an effective strategy for reaching zero dose children, increasing routine immunization coverage, and improving equity, but may need to be followed-up with strong routine services to have long-lasting impact. Further work is needed to understand the sustainability and cost-effectiveness of this approach compared with other methods for increasing coverage, particularly for the unreached populations.

## Data Availability

All data analyzed in this manuscript are publicly available. Outcome data are available from India's Health & Medical Information System (HMIS) and covariate data are available from the Demographic and Health Surveys (DHS).

https://nrhm-mis.nic.in/SitePages/Home.aspx

## Acknowledgements

This study was funded by the Bill & Melinda Gates Foundation. The authors thank participants at the International Heath Economics Association World Congress in July 2019 for their helpful input.

